# Beyond Identifier Matching: An Empirical Characterization of Failure Modes in Biomedical Knowledge Graph Integration

**DOI:** 10.64898/2026.05.26.26354182

**Authors:** Shiyue Hu, He Cheng, Lucas Gillenwater, Keenan Manpearl, Aishwarya Mandava, Yifan Wang, Milton Pividori, Barbara Stranger, Arjun Krishnan, Casey S. Greene, Yanjun Gao

## Abstract

**Objective:** Biomedical knowledge graphs (KGs) such as PrimeKG, Hetionet, UMLS, and PharmGKB are increasingly used as the substrate for downstream machine-learning, retrieval-augmented generation, drug-repurposing, and electronic health record (EHR) augmentation pipelines. The dominant assumption in published work is that integrating two or more such KGs is a tractable engineering step solved by identifier (ID) matching. This paper interrogates that assumption empirically. We quantify how much concept overlap survives realistic alignment, and we characterize the new failure modes introduced by the methods that practitioners reach for when ID matching is insufficient.

**Materials and Methods:** We compared four widely used biomedical KGs (PrimeKG, Hetionet v1.0, the full UMLS Metathesaurus, and PharmGKB) across eleven node types using a tiered alignment pipeline: (1) direct ID matching for nodes sharing a primary vocabulary; (2) cross-ontology bridging using standard mappings (e.g., MONDO↔DOID, HPO↔UMLS, HPO↔UMLS↔MeSH for side effects, NCBI Gene↔HGNC↔UMLS, UBERON↔FMA/SNOMEDCT_US/NCI/MeSH for anatomy); (3) ClinicalBERT cosine-similarity grouping at threshold ≥ 0.98 for over-segmented disease nodes, with a deterministic suffix-stripping canonicalizer; (4) exact name matching for ontology-poor types (anatomy, REACTOME pathways); and (5) embedding-based fuzzy matching with UMLS lookup (SapBERT and ClinicalBERT) for free-text microbiome concepts. We applied the pipeline to a 698-concept gut-microbiome benchmark spanning taxa, pathways, and disease labels, validated grouping decisions against the curated SSSOM mappings released by the MONDO project, and audited the ClinicalBERT consolidation against five clinical-genetics case studies drawn from the literature.

**Results:** Per-type pairwise coverage was strikingly asymmetric. Genes/proteins and the three Gene Ontology categories aligned cleanly across PrimeKG and Hetionet (mutual coverage 94–99%), but disease overlap was sparse: only 0.7% of PrimeKG individual disease nodes mapped to Hetionet, rising to 2.0% after MONDO grouping (versus 78.7% and 18.4% from the Hetionet side). PrimeKG-to-UMLS coverage spanned 100% (effect/phenotype via HPO) down to 20.8% (REACTOME pathways), with drugs at 73.7% and anatomy at 58.8%. PrimeKG-to-PharmGKB drug coverage required up to two bridging hops (DrugBank → UMLS → RxNorm/ATC/MeSH). Bigger was not uniformly more complete: on a 698-concept microbiome drug benchmark, Hetionet missed 0 concepts while PrimeKG missed 16. ClinicalBERT-based grouping consolidated 22,205 raw MONDO disease nodes into 17,080 groups but introduced three reproducible failure modes documented in case studies: (i) peer over-merging: for example, all 22 osteogenesis imperfecta subtypes collapsed into a single node despite distinct severity classes; (ii) parent-child collapse: e.g. acute myeloid leukemia merged with myeloid leukemia, erasing the acute/chronic distinction that drives clinical management; and (iii) lexical false positives: neurofibromatosis and schwannomatosis grouped together despite cellular-pathology differences.

**Discussion:** Identifier matching alone is a weak baseline for biomedical KG integration. Cross-ontology bridges and embedding-based consolidation expand coverage but do so at the cost of clinically meaningful resolution, and the resulting failures are systematic rather than random. Reporting only aggregate coverage statistics obscures these losses, which propagate silently into downstream tasks.

**Conclusion:** We provide reusable per-type coverage tables, a taxonomy of three integration failure modes, and concrete recommendations for downstream studies that depend on a unified biomedical KG. We argue that future KG integration work should report per-type coverage and per-cluster confidence rather than aggregate match rates.

## 1 BACKGROUND AND SIGNIFICANCE

Large-scale biomedical knowledge graphs have become a default substrate for translational machine learning. PrimeKG (Chandak et al., 2023) aggregates 20 primary resources into a multimodal disease-centric graph with text features and indication/contraindication edges. Hetionet (Himmelstein et al., 2017) integrates 29 sources into a biomedical knowledge graph with carefully curated edge semantics and provenance. UMLS (Bodenreider, 2004) supplies the most comprehensive controlled vocabulary in clinical informatics. PharmGKB (Whirl-Carrillo et al., 2021) is the de facto reference for pharmacogenomic relationships. Increasingly, papers do not pick one of these resources; they pick two or more and merge them, to widen drug coverage, to add side-effect signal, to connect microbial taxa to host phenotypes, or to ground a language model in structured biomedical relationships (Friedrichs, 2021).

In published methodology, this merging step is rarely the focus. It is often treated as a technical harmonization step based on shared identifiers and then regarded as resolved (Friedrichs, 2021). Reviewers, in turn, treat it as a low-risk engineering decision. This convention is at odds with what practitioners encounter when they actually attempt the merge.

Three structural mismatches make biomedical KG integration substantially harder than it is portrayed. First is granularity asymmetry: PrimeKG carries 22,205 fine-grained MONDO (Vasilevsky et al., 2022) disease nodes (collapsed to 17,080 by an internal grouping step), while Hetionet carries 137 hand-curated diseases drawn from the Disease Ontology (Schriml et al., 2022). Even after grouping, PrimeKG still contains far more disease nodes than Hetionet, differing by more than two orders of magnitude. As a result, a substantial fraction of PrimeKG diseases have no direct Hetionet counterpart, so analyses that depend on cross-KG disease alignment are necessarily constrained by the coarser Hetionet disease resolution. Second is version and scope drift: PrimeKG draws on DrugBank v5.1.8 (2021) (Wishart et al., 2018), with 7,957 drugs spanning approved, experimental, and investigational compounds, whereas Hetionet draws on DrugBank v4.2 (∼ 2015), restricted to 1,552 approved small molecules. Drugs introduced after 2015 (tirzepatide is one example) are simply absent from Hetionet, regardless of whether the merging code matched their identifiers. Third is competing node-typing decisions: PrimeKG collapses disease phenotypes and drug side effects into a single 15,311-node “effect/phenotype” type using HPO (Köhler et al., 2021) identifiers, while Hetionet keeps two separate types (Side Effect, with 5,734 UMLS-keyed nodes; Symptom, with 438 MeSH-keyed (Lipscomb, 2000) nodes) and provides no explicit phenotype type. The same clinical concept, for example nausea, exists as one node in PrimeKG (HP:0002018) and as two separate nodes in Hetionet.

Tooling for ontology alignment has matured considerably. Standardized mapping formats (SSSOM) (Matentzoglu et al., 2022), shared biomedical graph schemas such as the Biolink Model (Unni et al., 2022), curated cross-references (MONDO and HPO release explicit xrefs to DOID, UMLS, MeSH, and others), ontology-aware aligners (OAK (Mungall et al., 2024), the SSSOM toolkit), and NCATS Translator infrastructure for node normalization and reasoning (Fecho et al., 2022) make mechanical mapping reproducible. Related graph-transformation tools such as OWL-NETS (Callahan et al., 2018) further support concept abstraction and ontology-to-network conversion. Embedding-based normalization with biomedical language models (ClinicalBERT (Alsentzer et al., 2019), SapBERT (Liu et al., 2021), BioBERT (Lee et al., 2020)) extends the reach of these tools to free-text concepts that have no clean ontology presence. Each of these methods has been independently evaluated on its own benchmark. None, to our knowledge, has been audited end-to-end as the integration layer between two flagship biomedical KGs.

This paper supplies that audit. We measure how much concept overlap actually materializes between PrimeKG, Hetionet, UMLS, and PharmGKB under realistic alignment, decompose the result by node type, and characterize the systematic failures introduced by the alignment methods themselves.

## 2 OBJECTIVE

We pursue three objectives. First, to quantify pairwise concept coverage among PrimeKG, Hetionet, UMLS, and PharmGKB at the node-type level, separating matches obtained by shared identifiers from matches obtained through cross-vocabulary ontology mappings and from embedding-based consolidation. Second, to characterize the new failure modes that the methods used to expand coverage beyond ID matching introduce, using a case-study design grounded in the clinical-genetics literature. Third, to translate these results to a downstream task, namely a 698-concept gut-microbiome benchmark spanning taxa, pathways, and disease labels, and to show how integration choices propagate to use-case coverage.

## 3 DATASETS AND METHODS

### 3.1 Knowledge graphs

PrimeKG (Chandak et al., 2023) is a disease-centric multimodal graph with eleven node types. We used the public release without modification. Disease nodes are keyed to MONDO (Vasilevsky et al., 2022); the release ships 22,205 raw MONDO disease nodes, which we additionally consolidated to 17,080 grouped disease nodes using the procedure described in Sec. 3.3. Drug nodes are keyed to DrugBank v5.1.8 (Wishart et al., 2018) and total 7,957. Effect/phenotype nodes are keyed to HPO (Köhler et al., 2021) and total 15,311.

Hetionet v1.0 (Himmelstein et al., 2017) is a pure-graph integration of 29 sources with edge metadata and provenance. We used the publicly available Hetionet v1.0 Neo4j database distribution. Disease nodes (137) are keyed to the Disease Ontology (DOID) (Schriml et al., 2022); compound nodes (1,552) are keyed to DrugBank v4.2 (Wishart et al., 2018); side-effect nodes (5,734) are keyed to UMLS (Bodenreider, 2004); symptom nodes (438) are keyed to MeSH (Lipscomb, 2000); gene nodes (20,945) are keyed to NCBI Gene (Brown et al., 2015).

UMLS 2024AA Metathesaurus (Bodenreider, 2004) served as the cross-vocabulary bridge for ID conversions and as a reference for free-text concept matching. We used the full release and SapBERT-encoded UMLS index (Liu et al., 2021) for fuzzy lookup.

PharmGKB (Whirl-Carrillo et al., 2021) was used in the comparison of pharmacogenomic coverage against PrimeKG. We used the public release; gene, drug, phenotype, and chemical nodes were considered.

KG-Microbe (Joachimiak et al., 2024) was used as a microbiome-specific reference KG for the downstream benchmark of Sec. 3.5. We used the public release without modification and evaluated its coverage of microbial taxa and microbial pathway concepts. In contrast to PrimeKG, Hetionet, UMLS, and PharmGKB, KG-Microbe was included specifically to assess microbiome-domain coverage rather than host-disease or pharmacogenomic coverage.

### 3.2 Alignment pipeline

We applied a five-tier alignment pipeline. Tiers were applied per node-type pair and reported separately, since the appropriate tier depends on whether the source and target vocabularies are shared, related, or disjoint.

- **Tier 1, direct identifier matching:** applied where the two graphs share a primary vocabulary. NCBI Gene IDs (Brown et al., 2015) for gene/protein, GO IDs (Ashburner et al., 2000; The Gene Ontology Consortium, 2023) for the three Gene Ontology categories, DrugBank IDs (Wishart et al., 2018) for drug/compound.
- **Tier 2, cross-ontology bridging:** applied where the two graphs use related but distinct vocabularies and a standard cross-reference exists. We used MONDO↔DOID for disease (one-step) (Schriml et al., 2022; Vasilevsky et al., 2022), HPO→UMLS for the effect/phenotype symptom alignment (one-step), HPO→UMLS→MeSH for the effect/phenotype↔side-effect alignment (two-step), NCBI Gene→HGNC→UMLS (Seal et al., 2023) for gene-level UMLS coverage (two-step), and UBERON→*{*FMA, SNOMEDCT US, NCI, MeSH*}*→UMLS via five UBERON xref bridges for anatomy (Donnelly, 2006; Mungall et al., 2012; Rosse and Mejino, 2003; Sioutos et al., 2007).
- **Tier 3, exact name matching:** applied to ontology-poor types. REACTOME pathway names (Gillespie et al., 2022) were normalized (lowercased, whitespace collapsed) and matched against UMLS preferred terms; anatomy fell back to name matching when xref bridges produced no candidate.
- **Tier 4, ClinicalBERT cosine grouping:** applied to consolidate over-segmented MONDO disease nodes within PrimeKG before merge with Hetionet (Alsentzer et al., 2019). The procedure has two stages. Stage 1 is a deterministic suffix-stripping canonicalizer: tokenize on whitespace; iteratively drop trailing tokens that are numeric, two characters or fewer, or one of *{*type, variant, form, subtype}. For example, “Charcot-Marie-Tooth disease type 1A” reduces to “Charcot-Marie-Tooth disease”; “Charcot-Marie-Tooth disease type 2” reduces to the same canonical form. Stage 2 computes pairwise ClinicalBERT [CLS] cosine similarities among canonical forms and merges any pair with similarity ≥ 0.98.
- **Tier 5, embedding-based fuzzy matching:** applied to free-text microbiome concepts (taxa names, MetaCyc pathway labels (Caspi et al., 2020)) for which no standard identifier exists. We tokenized the source label, generated a query string by removing internal codes (e.g., “PWY-5534: propylene degradation” → “propylene degradation”), encoded it with SapBERT (Liu et al., 2021), and retrieved the nearest UMLS concept by cosine similarity, accepting matches at sim ≥ 0.95 after manual spot-check tuning.

### 3.3 ClinicalBERT disease grouping in PrimeKG

PrimeKG ingests each MONDO disease as a separate node, including subtypes that differ only by numeric or letter suffixes. This produces severe over-segmentation: osteogenesis imperfecta appears as 22 separate nodes (Types 1-21 plus several historical labels), Usher syndrome as 19 nodes, limb-girdle muscular dystrophy as 34 nodes. To make PrimeKG/Hetionet disease alignment tractable, we apply the Tier 4 procedure described above. We report two coverage rows: “disease (individual)” before grouping and “disease (grouped)” after grouping.

This consolidation is consequential. We therefore audited it against a curated SSSOM mapping (Matentzoglu et al., 2022) released by the MONDO project and against five clinical-genetics case studies (osteogenesis imperfecta, Usher syndrome, limb-girdle muscular dystrophy, MODY, and mucopolysaccharidoses) drawn from the literature. We classified each grouping decision into one of three outcomes (correct merge, peer over-merge, or parent-child collapse) and additionally examined low-similarity pairs that were not merged but exhibit lexical similarity, to bound false-positive rates if the threshold were lowered.

### 3.4 Reporting metrics

For each ordered node-type pair (source → target), we report the count of source nodes that resolve to at least one target node, the percentage of source nodes covered, the percentage of target nodes covered, and the maximum number of conversion steps required by the matching tier used. A pair may have multiple alignment paths; we report the maximum-coverage path and note alternatives in supplementary tables.

### 3.5 Downstream benchmark: gut microbiome concepts

We constructed a 698-concept benchmark drawn from a published 4,398-sample gut-microbiome study (Pasolli et al., 2017), retaining concepts present in more than 10% of samples. The benchmark spans three concept families: 1,211 taxa names, 619 MetaCyc pathway labels (Caspi et al., 2020), and 37 disease labels (the 37 includes composite labels such as “HF;CAD;T2D”, expanded into their atomic components for matching). For each KG we computed the fraction of benchmark concepts that resolve to at least one node, using the alignment tiers of Sec. 3.2 with embedding-based fuzzy matching enabled for taxa and pathways. We additionally computed the disease-restricted overlap, since the downstream taxon/function analysis is conditioned on disease state.

### 3.6 Reproducibility

All alignment scripts, the SapBERT/ClinicalBERT inference code, the UMLS index, the per-type coverage tables, the consolidated PrimeKG disease grouping, and the case-study annotations will be released under a permissive license at the time of publication. KG-specific licensing terms (notably for UMLS (Bodenreider, 2004) and DrugBank (Wishart et al., 2018)) prevent us from redistributing the source graphs, but the alignment pipeline operates on the public releases without modification.

## 4 RESULTS

We organize the results around the alignment tier that produces them. Each subsection embeds the table that anchors its claim. Section 4.6 reports the failure modes that arise when alignment relies on embedding-based consolidation; the failure modes themselves and their downstream implications are dissected per case in Sec. 5.

### 4.1 Direct ID matching reveals dramatic asymmetry between PrimeKG and Hetionet

Across eleven node-type pairs, mutual coverage is high for shared-vocabulary categories (genes, the three Gene Ontology branches) but collapses sharply for disease, anatomy, pathway, and effect/phenotype. The asymmetry is the more important finding: percentage coverage on the source side and the target side disagree by more than one order of magnitude in five of the eleven rows. Reporting either side alone gives a misleading summary of the merge.

- Headline: gene/protein contributes 19,003 overlapping nodes (68.8% of PrimeKG, 99.3% of Hetionet); the three GO categories all sit at 94% to 98% on the Hetionet side.
- Counter-headline: only 0.7% of PrimeKG individual disease nodes map to Hetionet (vs. 78.7% from the Hetionet side); after MONDO grouping the figures are 2.0% / 18.4%.
- Anatomy and pathway require name matching (Tier 3), so ID matching captures zero overlap for these types.

### 4.2 Cross-ontology bridges expand effect/phenotype coverage but split it across two Hetionet types

PrimeKG collapses disease phenotypes and drug side effects into a single “effect/phenotype” node type keyed to HPO (Köhler et al., 2021). Hetionet keeps two separate types, namely Symptom (MeSH) and SideEffect (UMLS). A single PrimeKG node can therefore have a counterpart in either Hetionet type, and the two bridges produce non-overlapping coverage.

- HPO→UMLS bridge (1 step): 271 PrimeKG nodes map to Hetionet Symptom (1.8% of PrimeKG, 65.3% of Hetionet).
- HPO → UMLS → MeSH bridge (2 steps): 1,161 PrimeKG nodes map to Hetionet SideEffect (7.6% of PrimeKG, 20.4% of Hetionet).
- The two bridges share PrimeKG nodes but diverge in target type. Any merge that picks one bridge silently drops the alternative typing.

### 4.3 PrimeKG-to-UMLS coverage ranges from 20.8% to 100% by node type

UMLS (Bodenreider, 2004) is widely used as a normalization target. Coverage of PrimeKG node types in the UMLS Metathesaurus ranges across nearly the full possible interval, depending on whether the source ontology is mirrored cleanly in UMLS.

- Strongest coverage: effect/phenotype 100.0% (HPO xrefs are essentially complete), the three GO categories 99.2% to 99.6%, gene/protein 99.5%.
- Weakest coverage: REACTOME pathway 20.8% (UMLS pathway content is sparse), anatomy 58.8% (UBERON is not mirrored cleanly), drug 73.7%.
- Practical reading: “normalize everything to UMLS” loses 79% of pathway concepts and 41% of anatomy concepts. A study that reports only the strongest-type figure (“99.5% UMLS coverage”) is hiding this.

### 4.4 PrimeKG-to-PharmGKB requires multi-step bridging via PubChem, RxNorm, ATC, or MeSH

PharmGKB (Whirl-Carrillo et al., 2021) is the standard pharmacogenomic reference. Coverage of PrimeKG against PharmGKB illustrates the cost of multi-hop bridging (Nelson et al., 2011; WHO Collaborating Centre for Drug Statistics Methodology, 2024): drug coverage starts at 27.5% with direct DrugBank-ID matching and expands as additional bridges are added, with each bridge widening coverage but introducing additional semantic drift.

- Genes align cleanly: 22,230 NCBI Gene IDs overlap directly (80.5% of PrimeKG, 89.3% of PharmGKB).
- Drugs do not: 27.5% direct match rises with bridges through UMLS → RxNorm/ATC/MeSH, but each additional hop opens the door to one-to-many ambiguity.
- Disease and phenotype overlap (1,404 nodes) is again strongly asymmetric, at 4.3% of PrimeKG and 86.8% of PharmGKB.

### 4.5 Bigger is not uniformly more complete: a microbiome counterexample

We tested a 698-concept microbiome benchmark against three reference KGs. The result contradicts the size-monotonicity heuristic: the smaller Hetionet covers more benchmark concepts than the larger PrimeKG.

Decomposing the same benchmark by concept family (taxa, pathway, disease) shows that no single KG is best on all three. The downstream choice between KGs cannot be made on aggregate size alone; it depends on which concept family dominates the task.

- Full UMLS captures 933/1,211 taxa and 40/619 pathways.
- PrimeKG captures 0 taxa and 0 pathways at the ID level, since the taxa/pathway content simply is not represented.
- KG-Microbe (Joachimiak et al., 2024) (microbe-specific) captures 1,138 taxa and 279 pathways but contributes no host-disease coverage.
- Implication: a microbiome study cannot be served by any single one of these KGs. A hybrid integration is required (revisited in Sec. 6).

### 4.6 ClinicalBERT consolidation introduces three reproducible failure modes

Tier 4 of the pipeline (ClinicalBERT cosine ≥ 0.98 with deterministic suffix stripping) (Alsentzer et al., 2019) consolidates 22,205 raw MONDO disease nodes in PrimeKG into 17,080 grouped nodes. The aggregate compression rate is favorable, but the case-by-case audit (*n* = 5 clinical-genetics conditions, plus follow-up sampling) reveals three reproducible failure modes. We name them, locate them in the data, and reserve their full dissection (mechanism, downstream implication, mitigation) for Sec. 5.

- Failure mode A (peer over-merging): subtypes that share a parent term are collapsed into a single grouped node when only the type identifier distinguishes them. Affects osteogenesis imperfecta (22→1), Usher syndrome (19→1), limb-girdle muscular dystrophy (34→1), MODY (≥ 15 → 1), mucopolysaccharidosis (Types 1-6 →1).
- Failure mode B (parent-child collapse): a child concept differing from its parent only by a clinical modifier (“acute”, “classic”, “cutaneous”, “juvenile”, “atrophicans”) is merged with the parent. Affects acute myeloid leukemia ↔ myeloid leukemia, classic ↔ general MSUD, cutaneous ↔ general melanoma, juvenile/infantile ↔ general nephropathic cystinosis, keratosis pilaris atrophicans ↔ keratosis pilaris.

Failure mode C (lexical false positives): two nodes are merged because their surface forms are similar despite distinct biology. Affects neurofibromatosis ↔ schwannomatosis (shared -omatosis suffix; distinct cellular pathology) and hypogonadotropic ↔ hypergonadotropic hypogonadism (single morpheme difference; opposite endocrine etiologies).

## 5 FAILURE MODES AND THEIR IMPLICATIONS

This section dissects each failure mode introduced in Sec. 4.6. For every mode we provide (i) the empirical evidence as a case-study mini-table, (ii) the mechanism that produces it, (iii) the downstream implication for tasks that consume the merged KG, and (iv) a candidate mitigation. The three modes are independent: any one of them is sufficient to compromise a downstream task that relies on subtype-level disease resolution.

### 5.1 Failure mode A: peer over-merging (suffix loss)

#### Evidence

Subtypes of a disease that share a parent term are collapsed into a single grouped node when the suffix-stripping canonicalizer removes the only token that distinguishes them. Five disease families illustrate the pattern; in every case the consolidated node erases a clinically relevant axis of variation.

#### Mechanism

The canonicalizer iteratively drops trailing tokens that are numeric, two characters or fewer, or in *{*type, variant, form, subtype*}*. For peer subtypes that differ only by a numeric type identifier (“Type 1A”, “Type 2”, …), this strips the only distinguishing token and reduces every peer to the same canonical form. ClinicalBERT cosine on identical strings is 1.0 by construction, so all peers are merged at the 0.98 threshold regardless of their underlying biology.

#### Downstream implication

- **Drug-repurposing models lose subtype-specific treatment signal**. MODY 2 responds to diet only; MODY 3 to sulfonylureas; MODY 5 to insulin (Urakami, 2019). A merged MODY node makes these distinctions invisible.
- **Severity-stratified prognosis is impossible to learn**. Osteogenesis imperfecta Type II is perinatally lethal; Type I is mild (Van Dijk and Sillence, 2014). A merged OI node gives the model a single label.
- **Enzyme-replacement therapy (ERT) recommendations become unsafe**. Idursulfase treats MPS Type II only; elosulfase alfa treats MPS Type IV only (Muenzer, 2011). A merged MPS node cannot support correct ERT selection.

#### Mitigation

- Preserve numeric type tokens during canonicalization unless an explicit MONDO subclass relation has already collapsed the subtype hierarchy.
- Treat the threshold-merged group as a candidate cluster, not a final node, and surface it for expert review when its constituents span more than *k* MONDO subclasses (e.g., *k* = 3).

### 5.2 Failure mode B: parent-child collapse

#### Evidence

A child concept that differs from its parent only by a clinical modifier (“acute”, “classic”, “cutaneous”, “juvenile”, “atrophicans”) is merged with the parent when the modifier token is dropped or down-weighted. The resulting node represents neither the parent nor the child accurately.

#### Mechanism

The canonicalizer does not have an explicit list of clinical modifiers, but downstream similarity collapses parent and modified-child forms because (i) ClinicalBERT cosine is dominated by content tokens shared between them and (ii) the modifier’s contribution to the [CLS] representation is small relative to the disease term itself. The result is reproducible across modifier classes (acuity, anatomical site, age of onset, severity).

### Downstream implication

- **Treatment recommendation becomes unsafe**. AML is a medical emergency; CML is a chronic disease treated with tyrosine kinase inhibitors (Arber et al., 2016). A merged “myeloid leukemia” node cannot route a patient correctly.
- **Pediatric vs. adult care pathways collapse**. Infantile nephropathic cystinosis presents at 6 to 12 months with severe Fanconi syndrome; juvenile cystinosis is later-onset and milder (Gahl et al., 2002). The grouped node loses the age-specific care pathway.
- **Triage models trained on the merged graph systematically under-prioritize the rare severe variant**, because the prevalence-weighted node is dominated by the common benign parent. For example, keratosis pilaris atrophicans is rare and scarring while keratosis pilaris is common and benign (Thomas and Khopkar, 2012), so the merged node behaves as the latter.

#### Mitigation

- Maintain the MONDO subclass relation explicitly: if A is a B in MONDO, never collapse A into B regardless of cosine similarity.
- Maintain a domain-curated list of clinical modifiers (acute, chronic, classic, congenital, juvenile, infantile, cutaneous, mucosal, uveal, atrophicans, …) that block merging when they distinguish two candidates.

### 5.3 Failure mode C: lexical false positives

#### Evidence

Two semantically distinct concepts are merged because their surface forms are similar. We observed this when a shared morphological suffix dominates the similarity (-omatosis) and when a single opposing morpheme is insufficiently weighted (hypo- vs. hyper-).

#### Mechanism

ClinicalBERT cosine on short clinical labels is dominated by surface form. Suffix-driven similarity (“-omatosis” tumors of any cellular origin) and minimal prefix differences (hypo- vs. hyper-) are both under-weighted relative to the larger token-overlap signal. The 0.98 threshold is permissive enough to admit these as valid merges.

#### Downstream implication

- **Cellular-pathology reasoning collapses**. Neurofibromas are mixed-cell tumors; schwannomas are pure Schwann cell (Zhu et al., 2002). Any task that links cell-type to disease (genetic testing, histology, targeted therapy) becomes incorrect.
- **Endocrine-axis reasoning inverts**. Hypogonadotropic hypogonadism is a pituitary disorder (low gonadotropins); hypergonadotropic hypogonadism is a gonadal disorder (high gonadotropins,
- gonadal failure) (Boehm et al., 2015). Diagnostic reasoning over the merged node points to the wrong organ system.
- **RAG/LLM grounding propagates the error**. If the merged node is retrieved as evidence, the language model receives semantically inconsistent context and may generate incorrect clinical text.

#### Mitigation

- Augment ClinicalBERT cosine with a morphological penalty: pairs that differ only by a known opposing prefix (hypo-/hyper-, pre-/post-, intra-/extra-, hypo-/normo-) are not merged at any threshold below an explicit override.
- For shared-suffix concepts (-omatosis, -opathy, -emia), require an additional cellular-class or organ-system token to match before merging.
- Report per-cluster confidence (e.g., minimum pairwise cosine within the cluster) so that downstream consumers can apply their own admission threshold.

### 5.4 Cross-cutting implication: integration is task-relative

The three failure modes share a structural property: each is acceptable for some downstream tasks and unacceptable for others. Peer over-merging is harmless if the task only needs a coarse disease label (e.g., epidemiologic counting) and lethal if it needs subtype-specific treatment selection. Parent-child collapse is harmless for an organ-level overview and lethal for triage. Lexical false positives are harmless for entity-frequency statistics and lethal for any reasoning task that consumes the node as a conceptual atom. A KG integration cannot be judged in isolation; it must be judged against the operational decisions it will support.

## 6 DISCUSSION

### 6.1 Identifier matching is necessary but insufficient

Aggregate cross-KG coverage statistics commonly reported in the literature (“19,003 genes overlap”) obscure type-specific coverage gaps that determine whether a downstream task is even feasible. The convention of describing integration in a single methods sentence makes it impossible for readers to judge how much of the downstream signal survives the merge. We recommend that biomedical-KG papers report per-type coverage with both source and target denominators, the alignment tier used per type, and the number of conversion steps required.

### 6.2 Bigger is not uniformly more complete

The microbiome counterexample (Tables 4 and 5) refutes the size-monotonicity heuristic that is frequently invoked to justify KG selection. PrimeKG, despite being substantially larger overall than Hetionet, misses 16 of 698 benchmark drug and disease concepts that Hetionet covers fully, and zero of the benchmark’s 1,211 taxa are represented in PrimeKG at the ID level. Coverage is task-relative and concept-family-relative. Selection should be benchmarked against the actual concepts the downstream task will encounter, not against aggregate node counts.

**Table 1.** Per-type concept overlap, PrimeKG vs. Hetionet. Tier denotes the alignment method required.

| PrimeKG type | Hetionet type | Overlap | PrimeKG (%) | Hetionet (%) | Tier |
| --- | --- | --- | --- | --- | --- |
| gene/protein | Gene | 19,003 | 68.8 | 99.3 | ID |
| drug | Compound | 1,520 | 19.1 | 98.8 | ID |
| biological_process | biological_process | 10,910 | 38.1 | 95.9 | ID |
| molecular_function | molecular_function | 2,718 | 24.3 | 94.2 | ID |
| cellular_component | cellular_component | 1,355 | 32.4 | 97.4 | ID |
| disease (individual) | Disease | 107 | 0.7 | 78.7 | Cross-ontology (MONDO→DOID) |
| disease (grouped) | Disease | 25 | 2.0 | 18.4 | Cross-ontology + ClinicalBERT |
| anatomy | Anatomy | 399 | 2.8 | 99.8 | Name match |
| pathway | Pathway | 1,113 | 44.2 | 61.1 | Name match |
| effect/phenotype | Symptom | 271 | 1.8 | 65.3 | Cross-ontology (HPO→UMLS) |
| effect/phenotype | SideEffect | 1,161 | 7.6 | 20.4 | Cross-ontology (HPO→UMLS→MeSH) |

**Table 2.** Coverage of PrimeKG node types in the UMLS Metathesaurus.

| Type | Source | Alignment bridge | Steps | in UMLS (%) |
| --- | --- | --- | --- | --- |
| gene/protein | NCBI | NCBI Gene → HGNC → UMLS | 2 | 99.5 |
| biological_process | GO | OBO xref | 1 | 99.5 |
| cellular_component | GO | OBO xref | 1 | 99.6 |
| molecular_function | GO | OBO xref | 1 | 99.2 |
| effect/phenotype | HPO | OBO xref | 1 | 100.0 |
| exposure | CTD | Direct MeSH match (Davis et al., 2023) | 0 | 99.3 |
| disease (individual) | MONDO | MONDO xref | 1 | 87.2 |
| drug | DrugBank | ID + name fallback | 0 | 73.7 |
| anatomy | UBERON | UBERON → CUI bridges | 1-2 | 58.8 |
| pathway | Reactome | Exact name match | 0 | 20.8 |

**Table 3.** Coverage of PrimeKG node types in PharmGKB.

| PrimeKG | PharmGKB | Overlap | PKG (%) | PGKB (%) | Best bridge | Steps |
| --- | --- | --- | --- | --- | --- | --- |
| gene/protein | genes | 22,230 | 80.5 | 89.3 | NCBI Gene direct | 0 |
| drug | drugs | 2,186 | 27.5 | 58.6 | DrugBank → UMLS → RxNorm/ATC/MeSH | 0-2 |
| disease + phenotype | phenotypes | 1,404 | 4.3 | 86.8 | MONDO/HPO → UMLS | 0-1 |
| exposure | chemicals | 28 | 3.4 | 0.5 | MeSH → DrugBank/PubChem | 0-2 |

**Table 4.** Microbiome benchmark (698 concepts): per-KG missing-concept counts.

| KG | Concepts not covered (/698) | Examples |
| --- | --- | --- |
| UMLS Diagnosis (microbiome ref) | 1 | WF10 (drug) |
| PrimeKG | 16 | Alendronate, Beclomethasone, Carbachol, Estropipate, L-Carnitine, L-DOPA, Norethindrone, Pamidronate, Penicillin V, Porfimer, Risedronate, Tiludronate, WF10, Zoledronate (all drugs); anemia, obesity (diseases) |
| Hetionet | 0 | n/a |

**Table 5.** Microbiome benchmark coverage by concept family per source KG.

| KG / source | Pathway (n=619) | Taxa (n=1,211) | Disease (n=37) | Notes |
| --- | --- | --- | --- | --- |
| Full UMLS (general) | 40 | 933 | 34 | Misses acute_diarrhoea, recipient_before, soil-transmitted helminthiasis |
| PrimeKG | 0 | 0 | $\leq 2$ | Misses healthy, recipient_before, few polys |
| Hetionet | 8 | 0 | 28 | n/a |
| KG-Microbe | 279 | 1,138 | n/a | Microbe-specific ontology coverage |

**Table 6.** Peer over-merging: representative cases drawn from the clinical-genetics literature.

| Disease family | MONDO subtypes | After grouping | Clinical structure that is lost |
| --- | --- | --- | --- |
| Osteogenesis imperfecta | 22 | 1 | Five-class severity grading (Type I to V): mild, lethal (perinatal), progressively deforming, moderate, special calcification (Van Dijk and Sillence, 2014) |
| Usher syndrome | 19 | 1 | USH1: profound congenital deafness with vestibular failure;<br>USH2: hearing aid-correctable, normal balance;<br>USH3: progressive hearing loss. Distinct molecular classes (motor proteins MYO7A vs. scaffold proteins USH1C/Harmonin) (Millán et al., 2011) |
| Limb-girdle muscular dystrophy | 34 | 1 | 2018 ENMC reclassification removes several PrimeKG-grouped subtypes from LGMD entirely (e.g., LGMD 2V is Pompe disease, LGMD 1B is Emery-Dreifuss) (Straub et al., 2018) |
| MODY | 13 | 1 | MODY 2 (GCK): diet only. MODY 3/1: sulfonylurea-sensitive. MODY 5: insulin-dependent. Treatment selection diverges by subtype (Urakami, 2019) |
| Mucopolysaccharidosis | 15 | 1 | Each type has a distinct enzyme deficiency and a type-specific enzyme replacement therapy (idursulfase for Type II, elosulfase alfa for Type IV) (Muenzer, 2011) |

**Table 7.** Parent-child collapse: cases where a clinically critical modifier is lost during consolidation.

| Parent concept | Child merged into parent | Clinical distinction erased |
| --- | --- | --- |
| myeloid leukemia (MONDO:11951) | acute myeloid leukemia (MONDO:11052) | AML is a medical emergency requiring immediate chemotherapy; chronic myeloid leukemia is managed with targeted therapy. Treatment-recommendation downstream tasks become unsafe (Arber et al., 2016) |
| maple syrup urine disease (MSUD) | classic MSUD | Classic MSUD presents with metabolic crisis and cerebral edema in the first days of life and requires emergency intervention; intermediate MSUD is far milder (Strauss et al., 2020) |
| melanoma | cutaneous melanoma | Melanoma comprises cutaneous, mucosal, and uveal subtypes with very different clinical management (Shain and Bastian, 2016) |
| nephropathic cystinosis | juvenile + infantile nephropathic cystinosis | Infantile form presents at 6 to 12 months with severe Fanconi syndrome; juvenile form is later-onset and milder (Gahl et al., 2002) |
| keratosis pilaris | keratosis pilaris atrophicans | Common keratosis pilaris is benign “chicken skin”; atrophicans is a rare variant causing permanent alopecia and scarring (Thomas and Khopkar, 2012) |

**Table 8.** Lexical false positives: surface-form similarity producing semantically wrong merges.

| Concept A | Concept B (merged with A) | Why the merge is wrong |
| --- | --- | --- |
| neurofibromatosis | schwannomatosis | Both end in -omatosis and share substantial lexical structure.<br>Pathologically distinct: neurofibromas are mixed-cell tumors, whereas schwannomas are composed almost entirely of Schwann cells (Zhu et al., 2002) |
| hypogonadotropic hypogonadism | hypergonadotropic hypogonadism | Differ by a single morpheme (hypo- vs. hyper-).<br>Represent opposite endocrine etiologies: pituitary deficiency vs. gonadal failure with elevated gonadotropins (Boehm et al., 2015) |

### 6.3 Implications for downstream tasks

- **Microbiome taxon-function discovery:** no single KG suffices. UMLS dominates on host-disease and taxa coverage; KG-Microbe (Joachimiak et al., 2024) dominates on microbial pathways; PrimeKG dominates on host disease-drug indication structure. A hybrid integration is required, with explicit provenance per concept family.
- **EHR pathology prediction:** the choice of KG materially affects which patient-subgraph paths are representable. A diabetes-to-retinopathy path traverses different edge types under PrimeKG (indication/contraindication) than under Hetionet (expression/upregulation), and the merged-disease failure modes of Sec. 5 directly determine whether subtype-specific complications can be modeled.
- **RAG/LLM grounding:** the emerging convention to “ground the LLM in a biomedical KG” inherits the chosen KG’s blind spots. The 16 drugs missing from PrimeKG are silently absent from any retrieval; concepts collapsed by the failure modes of Sec. 5 are returned as semantically inconsistent context.

### 6.4 Reporting recommendations for KG integration work

- Report per-type coverage with both source and target denominators.
- Report the alignment tier used per type (ID, cross-ontology, name match, embedding) and the bridges traversed.
- For embedding-based consolidation, report the threshold, the canonicalization rule, and a clinical-genetics case-study audit covering at least the three failure modes named in Sec. 5.
- Release per-cluster grouping decisions (and per-cluster confidence) rather than only the aggregate count.

## 7 LIMITATIONS

- Single-version snapshot: PrimeKG, Hetionet, UMLS 2024AA, and PharmGKB will continue to evolve; coverage figures are point-in-time.
- ClinicalBERT cosine threshold of 0.98 is one operating point; sensitivity to alternative thresholds is reported in supplementary materials, but the headline conclusions concern the qualitative structure of failure modes, not the exact threshold.
- Manual case studies are bounded to disease-type consolidation; analogous failure modes likely exist for drug and phenotype consolidation but are not characterized here.
- No large-scale human expert evaluation: the case studies draw on the published clinical-genetics literature, but a panel-based assessment would strengthen the failure-mode taxonomy.
- The microbiome benchmark is from a single cohort; coverage may differ on cohorts with different geographic or disease distributions.

## 8 CONCLUSION

Biomedical knowledge graph integration deserves the empirical rigor that the field already applies to model evaluation. We have shown that identifier matching captures only a fraction of true cross-KG overlap, that the methods used to bridge the gap introduce systematic and reproducible failure modes, and that no single KG is uniformly more complete than another on a realistic downstream benchmark. We provide per-type coverage tables, a taxonomy of three failure modes (peer over-merging, parent-child collapse, lexical false positives), and a set of reporting recommendations intended to make the integration layer of biomedical-KG papers as auditable as the model layer.

## Data Availability

All data produced in the present work are available online. The per-type coverage tables, consolidated PrimeKG disease groupings, ClinicalBERT/SapBERT inference code, UMLS alignment index, and case-study annotations will be released under a permissive open-source license upon publication. The source knowledge graphs (PrimeKG, Hetionet v1.0, UMLS 2024AA, PharmGKB, and KG-Microbe) are publicly available at their respective repositories; note that UMLS and DrugBank require free license registration and cannot be redistributed directl

## 8.1 Acknowledgment

This study was funded by the U.S. National Library of Medicine, National Institutes of Health, under award number R00LM014308.

*Acknowledgments*

